# Occupational fall incidence associated with heated tobacco product smoking and lifestyle behaviors: a nationwide cross-sectional study in Japan

**DOI:** 10.1101/2025.02.16.25321430

**Authors:** Saki Tsushima, Kazuhiko Watanabe, Sora Hirohashi, Tomohiro Yoshimi, Yoshihisa Fujino, Takahiro Tabuchi, Masayoshi Zaitsu

**Affiliations:** Center for Research of the Aging Workforce, University of Occupational and Environmental Health, Japan, Fukuoka, Japan; Department of Environmental Epidemiology, Institute of Industrial Ecological Sciences, University of Occupational and Environmental Health, Japan, Fukuoka, Japan; Department of Public Health, Tohoku University, Miyagi, Japan

**Author notes:** Corresponding author: Masayoshi Zaitsu, MD, PhD Director and Professor, Center for Research of the Aging Workforce, University of Occupational and Environmental Health, Japan, 1-1 Iseigaoka, Yahatanishi-ku, Kitakyushu, Fukuoka, 807-8555 Japan. These authors contributed equally (Saki Tsushima, Kazuhiko Watanabe).

**Keywords:** Occupational falls, bone fractures, heated tobacco product, smoking, workplace

## Abstract

**Objectives:** To identify the association between heated tobacco product (HTP) smoking and occupational falls, particularly combined with other lifestyle variables, including sleeping duration and comorbidities.

**Methods:** This web-based nationwide cross-sectional study (September–November 2023) involved 18,440 current workers (mean age, 43 years; women, 43.9%) in Japan. The primary outcome was occupational fall incidence in the past year. The secondary outcome was fall-related fractures. Smoking status was categorized as never, former, or current smokers; current smokers further classified into exclusive cigarette, exclusive HTP, and dual smokers (both cigarettes and HTPs). Other behavioral factors and covariates included alcohol consumption, sleep duration, physical activity, comorbidities, body mass index, hypnotics/anxiolytics use, and sociodemographic variables. Incidence ratios (IRs) and 95% confidence intervals (CIs) for occupational falls were estimated using multilevel Poisson regression with robust variance.

**Results:** Occupational falls and fall-related fractures were reported by 7.3% and 2.8% participants, respectively, in the past year. Younger workers were more likely to experience occupational falls. Compared with never smokers, current smokers had higher occupational fall incidence (IR = 1.36, 95% CI: 1.20–1.54), with exclusive HTP (IR = 1.78) and dual smokers (IR = 1.64) showing particularly elevated incidences. Similar patterns were observed for fall-related fractures. Elevated IRs were also observed for other lifestyle variables, including short sleep duration and diabetes, with the strongest associations found among younger workers.

**Conclusions:** Current smoking, particularly HTP use, and lifestyle behaviors were significantly associated with occupational falls in Japan. These findings highlight the importance of lifestyle modifications to prevent occupational falls.

**KEY MESSAGES:** *What is already known on this topic:* Individual lifestyle behaviors, including physical activity, sedentary behaviors, and sleep quality, are associated with the risk of falls. However, the association between the risk of falls and smoking habits, including heated tobacco product (HTP) use, which has emerged as a public health concern in Japan, remains poorly understood.

*What this study adds:* Current smoking, particularly HTP use, is associated with occupational falls.

*How this study might affect research, practice or policy:* Smoking cessation can be considered a modifiable behavior for occupational fall prevention. Further studies are needed to investigate the relationship between smoking and the incidence of falls.

## BACKGROUND

Falls are a critical public health concern worldwide, with an estimated 684,000 fall-related deaths and 37.3 million nonfatal but serious fall-related injuries occurring annually. [1] Measures to prevent falls are being promoted in the workplace. [2, 3] In Japan, where the workforce is aging rapidly, the rising number of occupational falls among older workers has become a concern for occupational safety. According to the latest data compiled annually by the Japanese Ministry of Health, Labour and Welfare, approximately 36,000 occupational falls resulting in at least 4 days of work absence were reported in 2023. [4]

To address this urgent issue, efforts in Japan have primarily focused on mitigating environmental and socioeconomic risks—two of the four major risk categories (i.e., biological, behavioral, environmental, and socioeconomic). [5] The government has encouraged strategies such as organization of fall-preventive environments and educating workers on health and safety. [6] However, strategies targeting behavioral risks, such as improving individual workers’ lifestyle habits, have received less emphasis, largely owing to a lack of workplace-specific evidence.

Individual lifestyle behaviors, including physical activity, sedentary behaviors, and sleep quality, are reportedly associated with the risk of falls in the workplace. [7–9] However, the association between risk of falls and certain lifestyle behaviors such as smoking—including the use of new types of tobacco (e.g., heated tobacco products [HTPs], including IQOS and Ploom TECH)—remain poorly understood among workers of all age groups (i.e., both older and young workers). [10] Furthermore, nationally representative data on overall occupational falls, including those without work absence, remain unavailable.

This study aimed to identify the incidence of occupational falls among Japanese workers of all age groups using a nationwide large-scale dataset. We sought to investigate the association between HTP smoking and occupational fall incidence, particularly in combination with lifestyle behaviors, in this population.

## METHODS

### Study design, data setting, and participants

This nationwide, cross-sectional study utilized data from the Japan COVID-19 and Society Internet Survey (JACSIS) (https://jacsis-study.jp/). The JACSIS dataset was derived from a pooled panel of an internet research agency (Rakuten Insight, Inc.), which includes approximately 2.3 million panelists across all 47 prefectures in Japan. Details regarding the JACSIS, including quality controls, national representativeness, and other policies for panelists, have been published previously. [11,12] Data for this study were collected in September and November 2023.

Initially, 25,332 participants aged 20–74[years were included. Of them, 6,892 individuals who were not actively employed (e.g., homemakers, students, and unemployed individuals) were excluded. The final analytical sample comprised 18,440 current workers (mean age ± standard deviation, 43 ± 14.3 years).

Informed consent was obtained electronically from all participants before completing the internet-based questionnaire. This study was conducted in accordance with the guidelines outlined in the Declaration of Helsinki, 1964, and was approved by the Ethics Committee of the University of Occupational and Environmental Health, Japan (No. R4-054).

### Outcome measurement: occupational falls

The primary outcome was the occurrence of workplace falls over the past year. Participants responded to the question, “Have you experienced a fall while at work in the past year?” with the following options:

1. Experienced a fall for the first time in the past 2 months,
2. Experienced a fall for the first time in the past year,
3. Experienced a fall in the past year but not for the first time,
4. Did not experience a fall in the past year but had experienced one more than a year ago,
5. Never experienced a fall.

Participants who selected options 1, 2, or 3 were categorized as having experienced a fall (“fallers”), whereas those who selected options 4 or 5 were categorized as “non-fallers.” Workers who selected options 3 or 4 were also identified as “previous fallers.” Falls during commuting were excluded to focus exclusively on workplace falls.

As the secondary outcome, fall-related fractures were identified by asking, “Have there been days when you were unable to work due to a fracture caused by a fall at work?” Individuals who experienced a fall without a fracture were excluded from the analysis. Additionally, supplementary data were collected on falls without injuries, falls with any type of injury, and near misses (almost fell).

### Exposure: Smoking and other covariates, including lifestyle behavioral factors

Based on smoking status, participants were categorized as never smokers, former smokers, or cigarettes or HTPs at least once but had quit smoking by the time of study initiation. Current smokers were further classified as exclusive cigarette smokers, exclusive HTP smokers, or dual smokers (using both cigarettes and HTPs). Available HTP brands during the study included Ploom Tech, Ploom X, IQOS, glo, and lil HYBRID. [11,13,14]

Other lifestyle behavioral factors included drinking habits (none, < 2 drinks/day, and ≥ 2 drinks/day; e.g., beer, Japanese sake, shochu, wine, and whisky), sleep duration (0–5 h, 6–9 h, ≥ 10 h, and unknown), and physical activity level (low, moderate, and high) based on the International Physical Activity Questionnaire Short Form (IPAQ-SF). [15] Additional covariates included comorbidities (hypertension, dyslipidemia, and diabetes), body mass index (BMI), and habitual use of hypnotics or anxiolytics.

Confounders included age, sex, educational level (≤ 12 years [high school] or ≥ 13 years [college or university]), occupation class and industry sector, equivalent annual household income (< 1.5 million JPY [approximately 15,000 USD], ≥ 1.5 million JPY, and unknown), and workplace size (≤ 49 workers, 50–999 workers, ≥ 1000 workers, and unknown). Moreover, to assess the ability of individuals to function at work, the Work Functioning Impairment Scale (WFun) was utilized. [16]

### Statistical analysis

Descriptive statistics were compared using the t-test for continuous variables or Chi-square test for categorical variables. Incidence ratios (IRs) and 95% confidence intervals (CIs) for workplace falls were calculated using multilevel Poisson regression with robust variance, accounting for clustering within prefectures. All analyses were adjusted for confounders Model 3, specific smoking types were included and adjusted for all other behavioral factors. Additionally, age-specific analyses were conducted for participants aged 15–39 years and 40–64 years; analyses for those aged 65–74 years were excluded due to insufficient outcomes.

For sensitivity analyses, we examined IRs for fall-related fractures. Additionally, we analyzed IRs after excluding previous fallers, given their high risk of recurrence as indicated by prior data (PR = 23.3, 95% CI: 20.4–26.6). The alpha level was set at 0.05, and all p-values were two-sided. Data were analyzed using STATA/SE 18.0 (StataCorp LLC, College Station, TX).

## RESULTS

Among the 18,440 current workers, 8,096 were women (43.9%), and 44.3%, 47.5%, and 8.2% were aged 20–39 years, 40–64 years, and 65–74 years, respectively. Of the total, 7.3% (n = 1,338) experienced occupational falls in the past year, and background characteristics differed significantly between non-fallers and fallers, except for educational level (Table 1). Estimated annual incidences across different types of falls were 3.4% for falls without injuries, 3.8% for falls with any injury, 2.8% for fractures, and 9.9% for near misses (Table 2). Younger individuals were more likely to experience occupational falls.

**Table 1.** Background characteristics of 18,440 workers.

| Characteristics |  | Total | Non-faller | Faller<sup>a</sup> |
| --- | --- | --- | --- | --- |
| N |  | 18,440 | 17,102 | 1,338 |
| Smoking status | Never | 10,486 (56.9%) | 9,863 (57.7%) | 623 (46.6%) |
|  | Former | 4,967 (26.9%) | 4,590 (26.8%) | 377 (28.2%) |
|  | Current | 2,987 (16.2%) | 2,649 (15.5%) | 338 (25.3%) |
| Smoking type | Never | 10,486 (56.9%) | 9,863 (57.7%) | 623 (46.6%) |
|  | Former | 4,967 (26.9%) | 4,590 (26.8%) | 377 (28.2%) |
|  | Cigarette only | 1,793 (9.7%) | 1,670 (9.8%) | 123 (9.2%) |
|  | HTP only | 667 (3.6%) | 542 (3.2%) | 125 (9.3%) |
|  | Dual | 527 (2.9%) | 437 (2.6%) | 90 (6.7%) |
| Habitual drinking | None | 7,633 (41.4%) | 7,128 (41.7%) | 505 (37.7%) |
|  | < 2 drinks/day | 7,553 (41.0%) | 7,003 (40.9%) | 550 (41.1%) |
|  | ≥ 2 drinks/day | 3,254 (17.6%) | 2,971 (17.4%) | 283 (21.2%) |
| IPAQ-SF level | Low | 11,642 (63.1%) | 10,795 (63.1%) | 847 (63.3%) |
|  | Moderate | 4,715 (25.6%) | 4,461 (26.1%) | 254 (19.0%) |
|  | High | 2,083 (11.3%) | 1,846 (10.8%) | 237 (17.7%) |
| Sleep duration | 0–5 h | 4,881 (26.5%) | 4,286 (25.1%) | 595 (44.5%) |
|  | 6–9 h | 13,056 (70.8%) | 12,371 (72.3%) | 685 (51.2%) |
|  | ≥ 10 h | 127 (0.7%) | 91 (0.5%) | 36 (2.7%) |
|  | Unknown | 376 (2.0%) | 354 (2.1%) | 22 (1.6%) |
| Hypertension |  | 2,871 (15.6%) | 2,485 (14.5%) | 386 (28.8%) |
| Dyslipidemia |  | 2,223 (12.1%) | 1,924 (11.3%) | 299 (22.3%) |
| Diabetes |  | 972 (5.3%) | 762 (4.5%) | 210 (15.7%) |
| Hypnotics/anxiolytics use |  | 1,354 (7.3%) | 1,178 (6.9%) | 176 (13.2%) |
| BMI, mean (SD) |  | 22.2 (6.6) | 22.2 (6.5) | 22.6 (7.2) |
| BMI category | < 18.5 | 2,571 (13.9%) | 2,397 (14.0%) | 174 (13.0%) |
|  | 18.5–24.9 | 12,625 (68.5%) | 11,735 (68.6%) | 890 (66.5%) |
|  | ≥ 25 | 3,244 (17.6%) | 2,970 (17.4%) | 274 (20.5%) |
| Women |  | 8,096 (43.9%) | 7,647 (44.7%) | 449 (33.6%) |
| Age, mean (SD) |  | 43.2 (14.3) | 43.4 (14.3) | 40.1 (13.7) |
| Age category | 20–39 years | 8,165 (44.3%) | 7,449 (43.6%) | 716 (53.5%) |
|  | 40–64 years | 8,756 (47.5%) | 8,201 (48.0%) | 555 (41.5%) |
|  | 65–74 years | 1,519 (8.2%) | 1,452 (8.5%) | 67 (5.0%) |
| College or greater |  | 14,260 (77.3%) | 13,253 (77.5%) | 1007 (75.3%) |
| Income | ≤ 150 million yen | 724 (3.9%) | 642 (3.8%) | 82 (6.1%) |
|  | > 150 million yen | 14,051 (76.2%) | 13,005 (76.0%) | 1,046 (78.2%) |
|  | Unknown | 3,665 (19.9%) | 3,455 (20.2%) | 210 (15.7%) |
| Industry sector | Blue collar | 4,913 (26.6%) | 4,504 (26.3%) | 409 (30.6%) |
|  | Service | 6,323 (34.3%) | 5,896 (34.5%) | 427 (31.9%) |
|  | White collar | 7,204 (39.1%) | 6,702 (39.2%) | 502 (37.5%) |
| Occupational class | Blue collar | 1,861 (10.1%) | 1,623 (9.5%) | 238 (17.8%) |
|  | Service | 8,005 (43.4%) | 7,465 (43.6%) | 540 (40.4%) |
|  | White collar | 5,286 (28.7%) | 4,930 (28.8%) | 356 (26.6%) |
|  | Others | 3,288 (17.8%) | 3,084 (18.0%) | 204 (15.2%) |
| Workplace size | ≤ 49 workers | 5,750 (31.2%) | 5,364 (31.4%) | 386 (28.8%) |
|  | 50–999 workers | 5,985 (32.5%) | 5,492 (32.1%) | 493 (36.8%) |
|  | ≥ 1000 workers | 4,934 (26.8%) | 4,595 (26.9%) | 339 (25.3%) |
|  | Unknown | 1,771 (9.6%) | 1,651 (9.7%) | 120 (9.0%) |
| WFun | 7–13 points | 9,997 (54.2%) | 9,606 (56.2%) | 391 (29.2%) |
|  | 14–20 points | 4,450 (24.1%) | 4,089 (23.9%) | 361 (27.0%) |
|  | 21–27 points | 2,775 (15.0%) | 2,340 (13.7%) | 435 (32.5%) |
|  | 28–35 points | 1,218 (6.6%) | 1,067 (6.2%) | 151 (11.3%) |
| Previous faller |  | 2,970 (16.1%) | 1,858 (10.9%) | 1,112 (83.1%) |

Abbreviations: HTP, heated tobacco product; IPAQ-SF, International Physical Activity
Questionnaire Short Form; WFun, Work Functioning Impairment Scale.
<sup>a</sup> All *p*-values < 0.05 except for educational level.

**Table 2.** Occupational fall incidence by age category.

| Age | Total workers, n | Overall falls |  |  | No injury |  |  | Injury |  |  | Fracture |  |  | Near miss |  |  |
| --- | --- | --- | --- | --- | --- | --- | --- | --- | --- | --- | --- | --- | --- | --- | --- | --- |
|  |  | N | % | Per 100,000 workers | N | N | N | N | % | Per 100,000 workers | N | % | Per 100,000 workers | N | % | Per 100,000 workers |
| Total | 18,440 | 1,338 | 7.3% | 7,256.0 | 636 | 3.4% | 3,449.0 | 702 | 3.8% | 3,806.9 | 508 | 2.8% | 2,754.9 | 1,833 | 9.9% | 9,940.3 |
| 5-year age category |  |  |  |  |  |  |  |  |  |  |  |  |  |  |  |  |
| 20–24 | 1,438 | 134 | 9.3% | 9,318.5 | 56 | 3.9% | 3,894.3 | 78 | 5.4% | 5,424.2 | 64 | 4.5% | 4,450.6 | 189 | 13.1% | 13,143.3 |
| 25–29 | 3,177 | 311 | 9.8% | 9,789.1 | 118 | 3.7% | 3,714.2 | 193 | 6.1% | 6,074.9 | 143 | 4.5% | 4,501.1 | 378 | 11.9% | 11,898.0 |
| 30–34 | 1,615 | 130 | 8.0% | 8,049.5 | 46 | 2.8% | 2,848.3 | 84 | 5.2% | 5,201.2 | 62 | 3.8% | 3,839.0 | 156 | 9.7% | 9,659.4 |
| 35–39 | 1,935 | 141 | 7.3% | 7,286.8 | 63 | 3.3% | 3,255.8 | 78 | 4.0% | 4,031.0 | 57 | 2.9% | 2,945.7 | 198 | 10.2% | 10,232.6 |
| 40–44 | 1,857 | 119 | 6.4% | 6,408.2 | 53 | 2.9% | 2,854.1 | 66 | 3.6% | 3,554.1 | 46 | 2.5% | 2,477.1 | 162 | 8.7% | 8,723.7 |
| 45–49 | 1,952 | 128 | 6.6% | 6,557.4 | 63 | 3.2% | 3,227.5 | 65 | 3.3% | 3,329.9 | 48 | 2.5% | 2,459.0 | 200 | 10.2% | 10,245.9 |
| 50–54 | 1,859 | 131 | 7.0% | 7,046.8 | 80 | 4.3% | 4,303.4 | 51 | 2.7% | 2,743.4 | 32 | 1.7% | 1,721.4 | 191 | 10.3% | 10,274.3 |
| 55–59 | 1,609 | 102 | 6.3% | 6,339.3 | 60 | 3.7% | 3,729.0 | 42 | 2.6% | 2,610.3 | 28 | 1.7% | 1,740.2 | 133 | 8.3% | 8,266.0 |
| 60–64 | 1,479 | 75 | 5.1% | 5,071.0 | 50 | 3.4% | 3,380.7 | 25 | 1.7% | 1,690.3 | 18 | 1.2% | 1,217.0 | 125 | 8.5% | 8,451.7 |
| 65–69 | 943 | 39 | 4.1% | 4,135.7 | 26 | 2.8% | 2,757.2 | 13 | 1.4% | 1,378.6 | 8 | 0.8% | 848.4 | 64 | 6.8% | 6,786.9 |
| 70–74 | 576 | 28 | 4.9% | 4,861.1 | 21 | 3.6% | 3,645.8 | 7 | 1.2% | 1,215.3 | 2 | 0.3% | 347.2 | 37 | 6.4% | 6,423.6 |
| 10-year age category |  |  |  |  |  |  |  |  |  |  |  |  |  |  |  |  |
| 20–29 | 4,615 | 445 | 9.6% | 9,642.5 | 174 | 3.8% | 3,770.3 | 271 | 5.9% | 5,872.2 | 207 | 4.5% | 4,485.4 | 567 | 12.3% | 12,286.0 |
| 30–39 | 3,550 | 271 | 7.6% | 7,633.8 | 109 | 3.1% | 3,070.4 | 162 | 4.6% | 4,563.4 | 119 | 3.4% | 3,352.1 | 354 | 10.0% | 9,971.8 |
| 40–49 | 3,809 | 247 | 6.5% | 6,484.6 | 116 | 3.0% | 3,045.4 | 131 | 3.4% | 3,439.2 | 94 | 2.5% | 2,467.8 | 362 | 9.5% | 9,503.8 |
| 50–59 | 3,468 | 233 | 6.7% | 6,718.6 | 140 | 4.0% | 4,036.9 | 93 | 2.7% | 2,681.7 | 60 | 1.7% | 1,730.1 | 324 | 9.3% | 9,342.6 |
| 60–69 | 2,422 | 114 | 4.7% | 4,706.9 | 76 | 3.1% | 3,137.9 | 38 | 1.6% | 1,569.0 | 26 | 1.1% | 1,073.5 | 189 | 7.8% | 7,803.5 |
| 70–74 | 576 | 28 | 4.9% | 4,861.1 | 21 | 3.6% | 3,645.8 | 7 | 1.2% | 1,215.3 | 2 | 0.3% | 347.2 | 37 | 6.4% | 6,423.6 |

Current smokers showed a significantly higher IR for occupational falls (IR = 1.36, 95% CI: 1.20–1.54), with exclusive HTP smokers and dual smokers exhibiting particularly elevated incidences (IR = 1.78 and 1.64, respectively) (Table 3). Other lifestyle and behavioral factors, including high physical activity levels, extreme sleep durations (0–5 h or ≥ 10 h), comorbidities, habitual use of hypnotics or anxiolytics, and severe WFun scores were also associated with higher incidences of occupational falls.

**Table 3.**
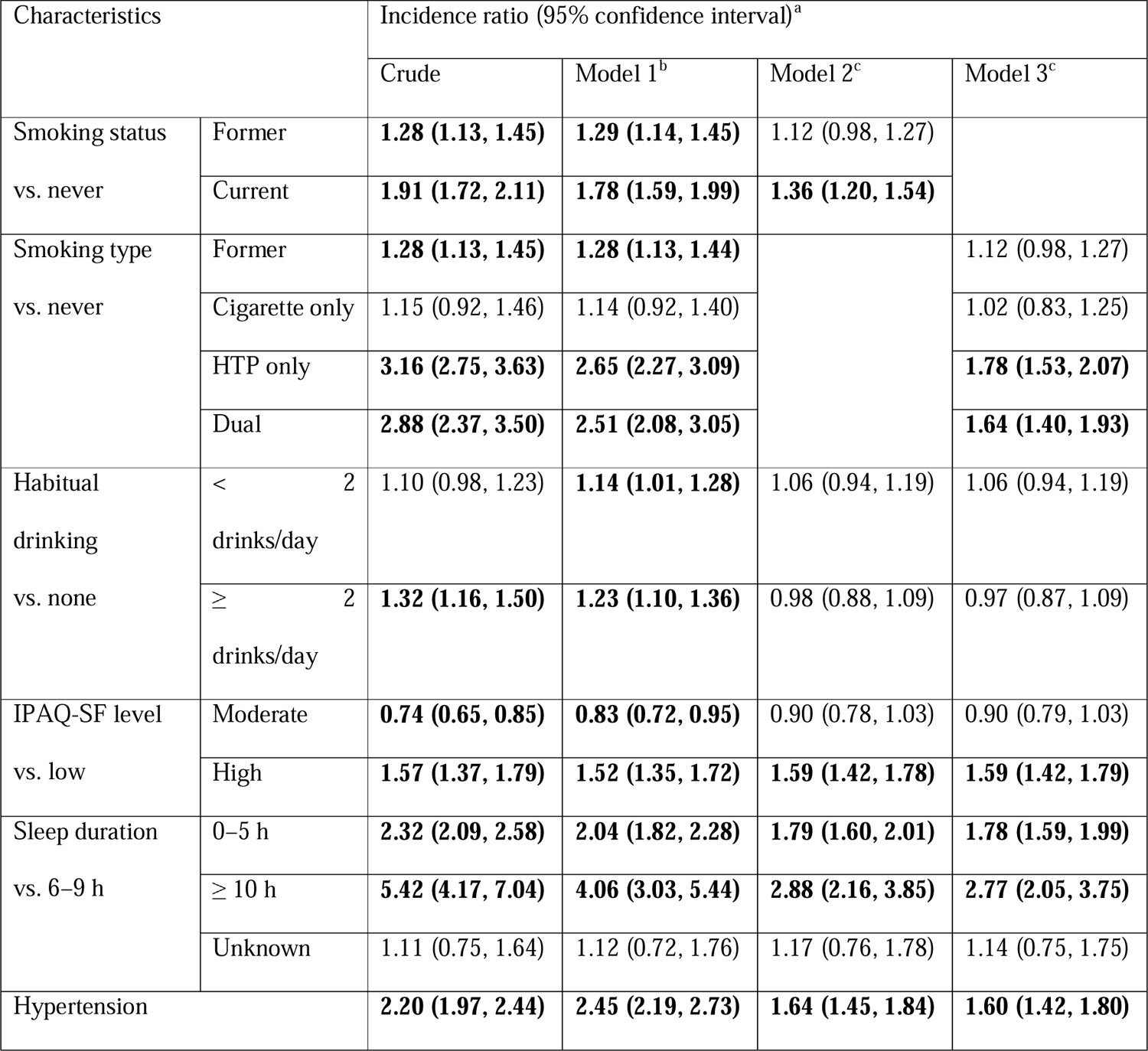
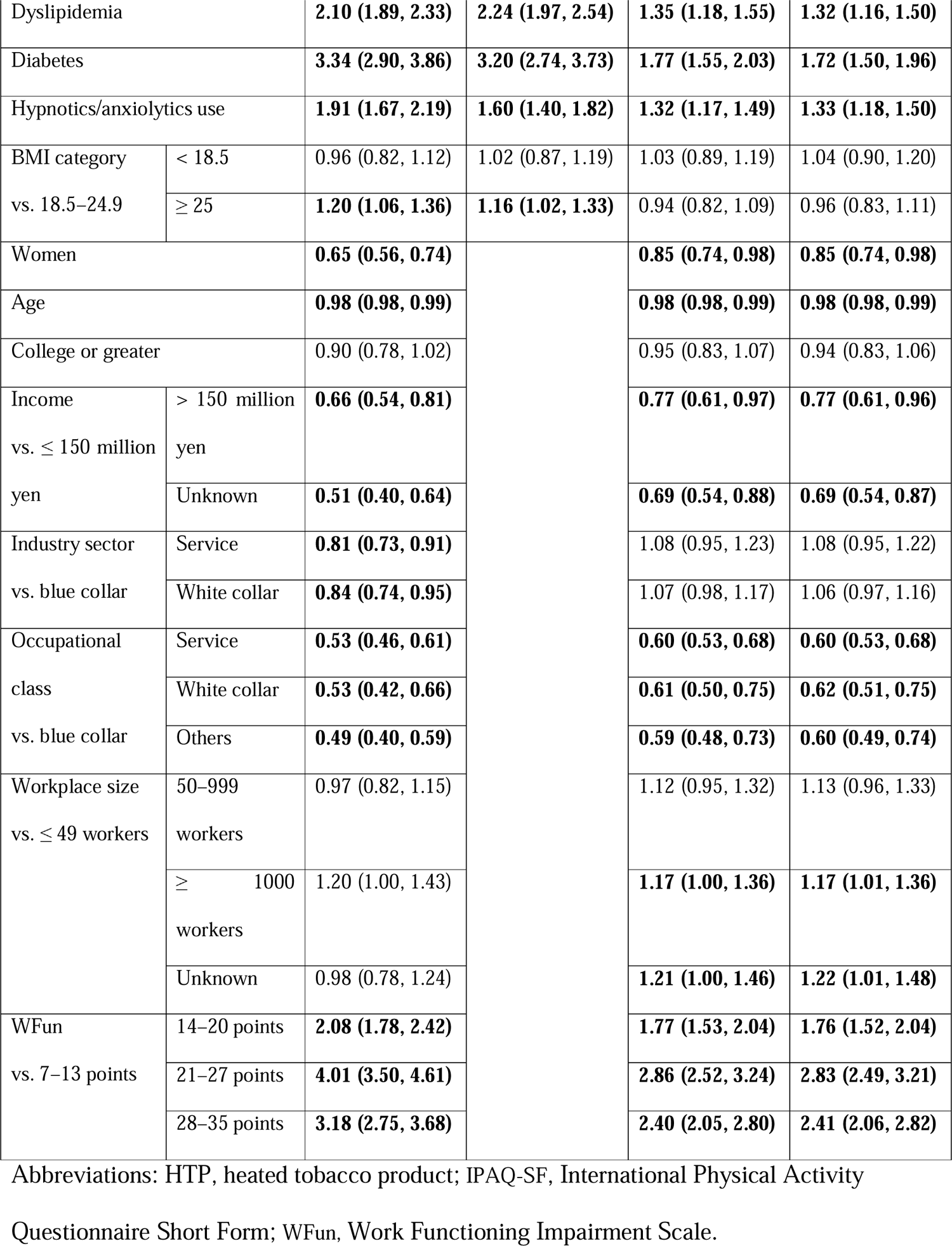
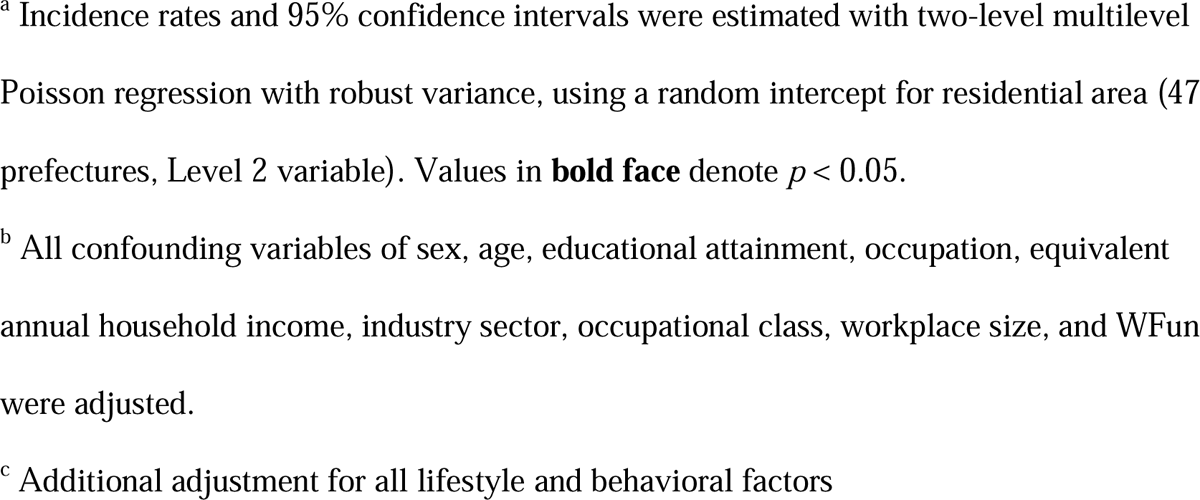
Incidence ratios of occupational fall across different smoking categories and other behavioral factors.

Subgroup and sensitivity analyses confirmed these patterns, with younger workers (aged 20– 39 years) showing the most pronounced associations (Figure 1).

**Figure 1.**
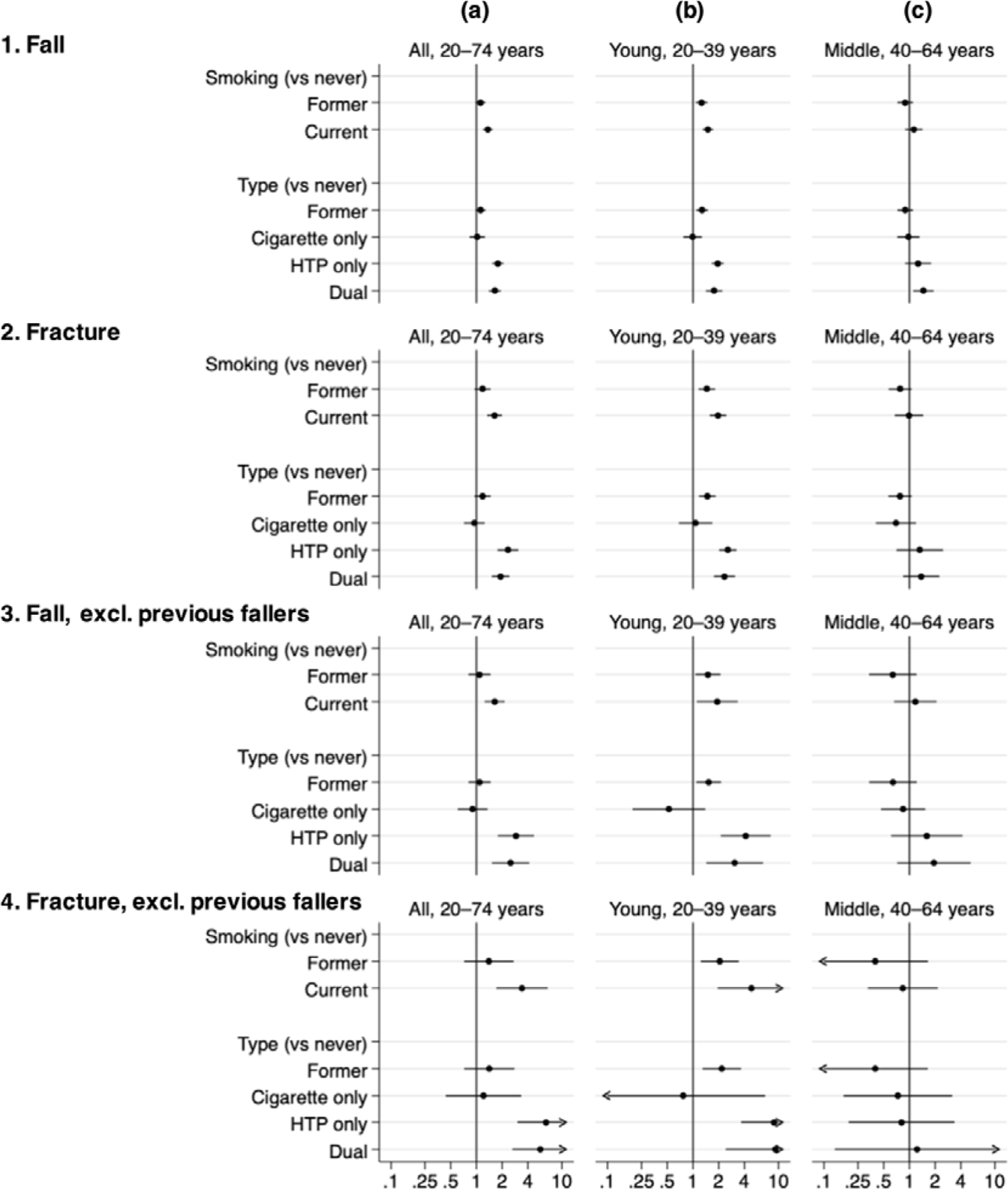
Incidence ratios of fall and fall-related fracture in each smoking category. The incidence ratios (circles) and 95% CIs (bars) of current smoking status/types, compared with never smokers, in different populations were estimated with two-level multilevel Poisson regression with robust variance. Random intercepts for residential area (47 educational attainment, occupation class, industry sector, equivalent annual household income, workplace size, and Work Functioning Impairment Scale) and other lifestyle and behavioral variables (drinking habits, weekly physical activity, hypertension, dyslipidemia, diabetes, body mass index, and habitual use of hypnotics or anxiolytics). The samples used in the analysis were as follows: (1) n = 18,440 (1,338 falls and 17,102 non-fallers), (2) n = 17,610 (508 fractures and 17,102 non-fallers), (3) n = 15,470 excluding previous fallers (226 falls and 15,244 non-fallers), and (4) n = 15,300 excluding previous fallers (56 fractures and 15,244 non-fallers). Abbreviations: HTP, heated tobacco product; CI, confidence interval.

## DISCUSSION

In this nationwide, cross-sectional study conducted in Japan, we found that various lifestyle and behavioral factors, including HTP smoking, were associated with occupational falls in the workplace. Compared with never smokers, current smokers were associated with a 1.3-fold higher incidence of occupational falls, with the most pronounced association observed among HTP smokers, who exhibited approximately a 1.8-fold higher incidence. A similar pattern was observed for falls resulting in fractures, particularly among younger workers.

Other lifestyle and behavioral factors, including physical activity, sleep durations, comorbidities, and individual functioning at work, were also associated with occupational falls. These findings highlight the potential importance of lifestyle modifications, including smoking cessation, in preventing occupational falls.

In Japan, HTP smoking has emerged as a public health concern, with approximately 40% occupational falls and smoking remains limited, and no previous studies have specifically investigated the impact of HTP smoking on occupational falls. Epidemiological studies have reported an increased risk of fall-related fractures among current smokers. [18] A recent Mendelian randomization study also identified a higher tendency of falling among current smokers. [19] Smokers exhibit decreased postural stability compared with non-smokers, [20] and nicotine exposure can affect vestibular functions. [21] Consequently, new tobacco products, such as HTPs, may also biologically contribute to an increased incidence of occupational falls.

In our study, individual comorbidities, such as hypertension and diabetes, medication use (e.g., hypnotics or anxiolytics), engaging in high levels of physical activity, and short sleep duration, were associated with higher incidences of occupational falls, which is consistent with findings from previous studies. [9,22–24] We also found an association between individual functional ability at work and occupational falls. Therefore, although occupational safety should primarily focus on mitigating environmental risks, strategies targeting lifestyle and behavioral risks may also play a crucial role in preventing occupational falls.

A notable finding in our study is that, even among younger workers, smoking behavior— particularly HTP use—was associated with occupational falls and fall-related fractures. Interestingly, more than half of the occupational falls observed in our study occurred in this younger age group. Although fall prevention strategies have traditionally focused on older individuals considered to be at higher risk, [4] the incidence of falls across different age groups, particularly among younger workers, has not been fully elucidated. Our findings may emphasize the importance of addressing occupational falls among younger workers.

Workplace fall prevention strategies should also target relatively younger age groups. Hence, encouraging healthy lifestyle behaviors could play a critical role in reducing falls, potentially serving as a nudge effect, which is the result of using nudging techniques to influence people to make certain choices such as changing their lifestyle behaviors based on other important concerns. [25]

Our study had some limitations. First, as this was a self-reported, cross-sectional study, causal inferences could not be established. Additionally, detailed information on smoking, such as smoking intensity and duration of smoking abstinence, was not available. Further studies are warranted to investigate the mechanisms and temporal relationship between smoking and fall incidence. Second, recall and reporting bias of HTP use could not be discarded, as suggested in a study on combustible cigarette and electronic cigarette smoking. [26] Third, although sensitivity analyses demonstrated similar patterns, the data on falls were self-reported, which may have introduced underestimation or misclassification.

Despite these limitations, to the best of our knowledge, this nationwide study is the first to specify the incidence of occupational falls by smoking type and to reveal the association between HTP smoking and other behavioral factors, and occupational falls in Japan. Although the impacts of smoking on various health outcomes are widely established, greater attention should be paid to occupational falls among current smokers, regardless of tobacco type, from the perspective of occupational safety.

In conclusion, occupational falls may occur across all age groups, and various lifestyle behaviors, particularly HTP smoking, may be associated with occupational falls. To better understand the observed associations, further studies are needed to identify the mechanisms by which smoking and lifestyle behaviors contribute to the increased incidence of occupational falls in the aging society.

## Data Availability

The data that support the findings of this study are restricted due to personal identification and privacy concerns. The research data can be obtained from the corresponding author upon reasonable request.

## Acknowledgments

We would like to thank Editage (www.editage.com) for English language editing.

## Author contributions

ST, KW, and MZ designed the study. MZ supervised the study. ST, KW, and MZ developed the methodology. ST, TT, and MZ created the dataset. ST and MZ analyzed the data. ST, KW, and MZ wrote the first draft of the manuscript. SH, TY, YF, and TT commented on the manuscript. All authors read and approved the final version.

## Ethics approval

The study was approved by the Ethics Committee of the University of Occupational and Environmental Health, Japan (no. R4-054).

